# Trends in Cardiac Arrest Outcomes & Management in Children with Cardiac Disease: An Analysis from the AHA Get With The Guidelines^®^-Resuscitation Registry

**DOI:** 10.1101/2023.04.24.23289073

**Authors:** Monique M. Gardner, Ryan W. Morgan, Ron Reeder, Kimia Ghaffari, Laura Ortmann, Tia Raymond, Javier J. Lasa, Jessica Fowler, Maya Dewan, Vinay Nadkarni, Robert A. Berg, Robert Sutton, Alexis Topjian, the American Heart Association’s Get With The Guidelines^®^-Resuscitation Investigators

**Affiliations:** Department of Anesthesiology and Critical Care Medicine, The Children’s Hospital of Philadelphia; Perelman School of Medicine at the University of Pennsylvania, Philadelphia Pennsylvania; Department of Pediatrics, University of Utah, Salt Lake City, Utah; Department of Pediatrics, University of Nebraska Medical Center, Children’s Hospital & Medical Center Omaha, Nebraska; Department of Pediatrics, Cardiac Critical Care, Medical City Children’s Hospital, Dallas, Texas; Divisions of Cardiology and Critical Care Medicine, Children’s Health, UT Southwestern Medical Center, Dallas, TX; Division of Critical Care Medicine, Department of Pediatrics, Cincinnati Children’s Hospital Medical Center, Cincinnati, Ohio

## Abstract

**Introduction:** Contemporary rates of survival after pediatric in-hospital cardiac arrest (IHCA) and trends in survival over the last 20 years have not been compared based on illness category. We hypothesized that survival to hospital discharge for surgical-cardiac category is higher than the non-cardiac category, and rates of survival after IHCA increased over time in all categories.

**Methods:** The AHA Get With The Guidelines^®^-Resuscitation registry was queried for index IHCA events in children <18 years of age from 2000-2021. Categories were surgical-cardiac (IHCA following cardiac surgery); medical-cardiac (IHCA in non-surgical cardiac disease); and non-cardiac (IHCA in patients without cardiac disease). The primary outcome was survival to hospital discharge. We compared eras 2000-2004, 2005-2009, 2010-2014, and 2015-2021 with mixed logistic regression models, including event year as a continuous predictor and site as a random effect.

**Results:** Of 17,696 index events, IHCA rates by illness category were: 18% surgical-cardiac, 18% medical-cardiac, and 64% non-cardiac. Surgical-cardiac category had the highest rate of survival to discharge compared to medical-cardiac and non-cardiac categories (56% vs. 43% vs. 46%; p<0.001). After controlling for age, location of event, and hospital size, the odds of survival were highest for surgical-cardiac category (aOR 1.28, 95% CI 1.16-1.40) and lower for medical-cardiac category (aOR 0.87, 0.80-0.95), compared to the non-cardiac category. Odds of survival increased for all illness categories from the 2000-2004 era to the 2015-2021 era. Rates of improvement differed among illness categories with medical-cardiac having the lowest increased odds per era. Surgical-cardiac patients had the highest rates of extracorporeal resuscitation (ECPR) (20% across the cohort), though the greatest increase in ECPR utilization was in the non-cardiac population (44% increased odds per era).

**Conclusions:** Over the last 20 years, both survival to hospital discharge and ECPR use has increased in all IHCA illness categories. Children with surgical-cardiac IHCA have higher odds of survival to hospital discharge compared to non-cardiac IHCA categories, whereas odds of survival were lowest with medical-cardiac IHCA.

## INTRODUCTION

There are more than 19,000 pediatric in-hospital cardiac arrests (IHCA) per year in the United States, occurring in 1-2% of hospitalized children per year^1, 2^. Children with underlying cardiac disease are a unique population and have a higher incidence of IHCA: 2-6% of all children in cardiac care units^3^. Overall survival rates after IHCA have increased over the last 20 years from 19% to 38% for pulseless events and from 57% to 66% for non-pulseless events^4^.

IHCA in patients with underlying cardiac disease, including congenital heart disease or pediatric heart failure, differs from arrests due to non-cardiac causes; in part because these children have high rates of surgery for congenital heart disease prior to arrest, have complex physiology impacting resuscitation^5, 6^, and are cared for in settings where extracorporeal cardiopulmonary resuscitation (ECPR) is more likely to be available and rapidly deployed^7^. Furthermore, while survival rates from all pediatric IHCA are approximately 43%^8^, children with underlying perioperative cardiac disease have higher survival rates, up to 62%^3^. Pediatric cardiac arrest studies usually combine patients regardless of their pre-existing conditions^1, 9–13^, thus few studies have specifically evaluated outcomes and management after pediatric IHCA in infants and children with heart disease^14, 15^. In 2011, Ortmann *et al.* described disparities in outcomes between illness categories demonstrating higher rates of survival to hospital discharge for surgical-cardiac disease compared to medical-cardiac or non-cardiac patients^14^.

Rates of ECPR utilization, survival from IHCA, and survival from IHCA in children with congenital heart disease have increased over time^16, 17^. However, these studies do not address how cardiac arrest management and outcomes differ between children with surgical-cardiac and medical-cardiac compared to non-cardiac disease and how outcomes have changed over time.

To address these gaps, we used the American Heart Association’s (AHA’s) Get With The Guidelines^®^-Resuscitation (GWTG®-R) registry of cardiac arrests^18^ to (1) compare contemporary IHCA management and outcomes in children with cardiac disease to those without cardiac disease, and (2) to describe the change over time in management and outcomes after IHCA in children with cardiac disease compared to those without cardiac disease. We hypothesized that (1) survival to hospital discharge for surgical-cardiac illness category is higher than the non-cardiac illness category, and (2) rates of survival after IHCA increased over time in all categories.

## METHODS

This is a retrospective study using data from the AHA GWTG®-R registry, an in-hospital resuscitation quality improvement registry using Utstein-style data reporting. Standardized data collection forms are used to report all patients, employees and visitors who require resuscitation in a hospital facility. Data collection includes information about the facility, patient demographics, pre-event data, cardiopulmonary resuscitation (CPR) event data, and outcome data. Data collection and integrity has been previously described^19–21^. Hospitals participating in the registry submit clinical information regarding the medical history, hospital care, and outcomes of consecutive patients hospitalized for cardiac arrest. IQVIA (Parsippany, New Jersey) serves as the data collection (through their Patient Management Tool – PMT™) and coordination center for the American Heart Association/American Stroke Association Get With The Guidelines® programs. This study was deemed exempt by the Children’s Hospital of Philadelphia IRB because it was considered non-human subjects’ research. All participating institutions were required to comply with local regulatory and privacy guidelines and, if required, to secure institutional review board approval. Because data were used primarily at the local site for quality improvement, sites were granted a waiver of informed consent under the common rule.

Patients were included for analysis if they were <18 years of age with an IHCA requiring chest compressions from 2000-2021. Only index events (the first cardiac arrest per subject per admission) were evaluated. Patients were excluded if they did not receive compressions, if resuscitation occurred in the delivery room, if they had an illness category of obstetric, trauma or other, or were a non-patient (visitor or employee) (**Figure 1**). We excluded patients from sites that had <10 eligible patients during the 20-year study period.

**Figure 1.**
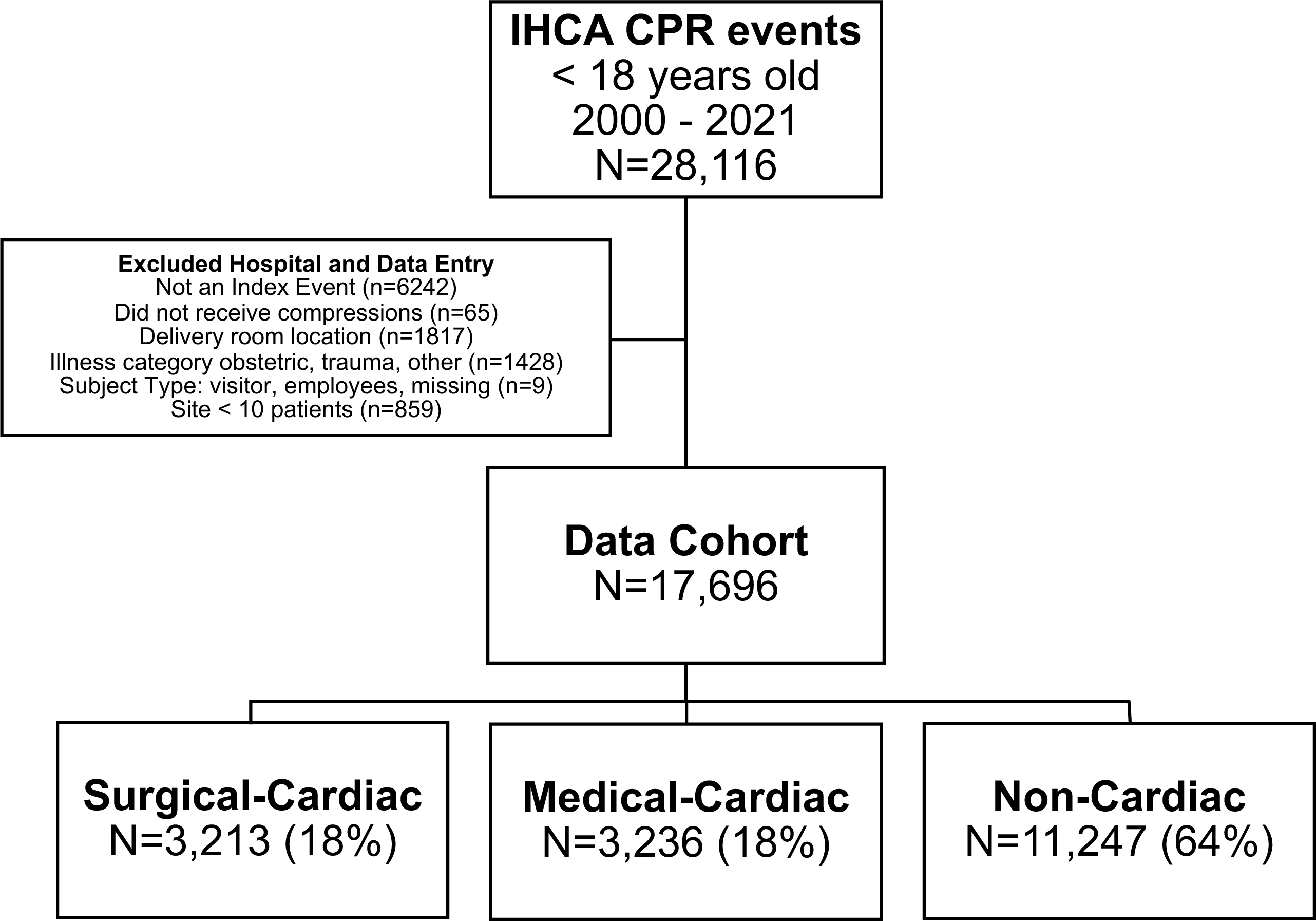
Consort diagram for subject inclusion from Get With The Guidelines^®^-Resuscitation.

GWTG®-R categorizes patients into illness categories which are selected upon data entry based on standard definitions: medical-cardiac, medical-noncardiac, surgical-cardiac, surgical- non-cardiac, obstetric, trauma and other^14^. Because our primary aim was to compare management and outcomes across illness categories, we created three illness category groups: (1) surgical-cardiac: patients who are had their index cardiac arrest during the same hospital admission as their cardiac surgery; (2) medical-cardiac: patients whose primary diagnosis is cardiovascular including congenital heart disease without cardiac surgery during the admission with a cardiac arrest; (3) and non-cardiac: medical-noncardiac patients (patients with medical illness that is non-cardiovascular in nature) and surgical-noncardiac patients (patients who are pre- or postoperative from a non-cardiac surgery during the same hospital admission as their cardiac arrest).

From 2005 to 2015, ‘newborn’ was a distinct illness category in the registry and was defined as a patient who was born on that admission or transferred from their birth hospital during same admission as the IHCA. Therefore, many patients with congenital heart disease on admission at birth would have been characterized as illness category “newborn” when they could have been “medical non-cardiac” or “surgical non-.cardiac.” The newborn illness category was removed in 2015 because newborn was determined not to be an illness category, but an age classification which was redundant with the other possible categories. Therefore, after 2015, patients could no longer be categorized into the “newborn” illness category and were categorized into medical-cardiac, medical-noncardiac, surgical-cardiac, surgical-noncardiac, trauma, or other. There were 2,200 eligible patients in the “newborn” illness category in our cohort prior to 2016. To avoid excluding these patients, and to prevent the analysis being driven by spurious trends due to the addition of the relabeled newborn illness category to the cohort starting in the year 2016, imputation was used to recategorize patients in the newborn illness category prior to 2016 as medical-cardiac, surgical-cardiac, or non-cardiac.

Imputation accomplishes two key objectives. First, imputation reduces bias by preventing the systematic exclusion of subjects with missing values, allowing all relevant subjects to be included in the analysis. Second, imputation makes full use of the data collected – even incomplete data – so that collected data isn’t wasted. Imputation was performed using multiple imputation by chained equations (MICE)^22^ in IVEware v0.2 (University of Michigan) for SAS. In this approach, regression models were used to estimate a set of likely values for the data that were missing. A sequence of stepwise regression models^23^ were created from pre-existing conditions, interventions at place at the time of the event, event and subject characteristics, interventions initiated during the event, event outcomes, and hospital characteristics variables. Variables used for imputation models and their rates of missingness prior to imputation are reported in **Supplemental Table 1**. A random component was incorporated to reflect the uncertainty about the missing values. This process was repeated to produce 50 datasets without missing values. The standard 9 iterations of the process were performed between datasets to allow model stabilization^24^. Analyses were then conducted on each of the 50 datasets individually. Results reported were based on averages across the 50 datasets, with confidence intervals and p-values (based on Rubin’s formula) that additionally incorporate the uncertainty due to missingness in the original dataset.

Pre-arrest and hospitalization characteristics and outcomes were summarized by illness category to characterize each patient population. Pre-existing conditions are comorbidities selected at time of data entry at the discretion of the site. Age was categorized as less than 1 month, 1 month to less than a year, 1 year to under 8 years, and 8 years to under 18 years. Cardiac arrest duration was truncated to 120 minutes of analysis due to concern for data entry errors, so any cardiac arrest duration of >120 minutes was recorded as 120 minutes. The same was done with the timing (minutes) to the first dose of epinephrine, truncating to 30 minutes, as doses administered after 30 minutes were rare. The receipt of any epinephrine, the number of doses of epinephrine given, and the time to first epinephrine dose (in minutes) were included in analysis.

The primary outcome was survival to hospital discharge. Secondary outcomes were event survival, defined as sustained return of circulation, either spontaneously (ROSC) or return of circulation with extracorporeal membrane oxygenation, ECMO (ROC). We reported survival with favorable neurologic outcome (defined as Pediatric Cerebral Performance [PCPC]^25^ score of 1 to 2 or no change from baseline) as an outcome, though this variable had a high percentage of missingness (28.9%) and was excluded in further analyses.

### Statistical Analysis

Data were summarized as counts and percentages for categorical variables and median and interquartile ranges for continuous variables. Associations with outcomes were assessed with the likelihood ratio test for binary and nominal characteristics and the Kruskal-Wallis test for ordinal and quantitative characteristics. These tests were chosen because they are readily adapted to multiply imputed data and account for the ordered or unordered nature of each characteristic, as appropriate.

We evaluated the association of illness category with survival to hospital discharge in a mixed logistic regression model, controlling for age, event location within the hospital, and hospital size. Trends over time were evaluated with mixed logistic regression models, including event year as a continuous predictor and site as a random effect. Site was included as a random effect to avoid spurious trends that may otherwise be identified due to sites with systematic differences (e.g., survival rate) starting or ending their participation at any point during the study period. A separate model was created for each illness category, and descriptive summaries were reported in 5-year increments, aligning with publication of AHA Pediatric Basic and Advanced Life Support (PALS) guidelines: 2000-2004, 2005-2009, 2010-2014, 2015-2021. Model estimates of survival to hospital discharge were additionally plotted over time. A separate mixed logistic regression model with era, illness category, and their interaction was created to determine if the trend in survival to hospital discharge was different between illness categories. Analyses were performed using SAS 9.4 (SAS Institute; Cary, NC). P-values were based on two-sided alternatives and considered significant when <0.05.

## RESULTS

The initial cohort included 28,116 events in children <18 years old between 2000 and 2021. After applying exclusion criteria, the evaluable cohort was 17,696 index events (**Figure 1**). Of the evaluable events, illness categories were surgical-cardiac (n=3,213; 18%), medical- cardiac (n=3,236;18%), and non-cardiac (n=11,247; 64%).

### Demographics and patient characteristics based on illness category

Surgical-cardiac patients were younger; more likely to have a cardiac arrest in a procedural area, a witnessed arrest, a pre-existing condition of hypotension/hypoperfusion, and less likely to have a cardiac arrest occur during a night or weekend compared to patients in medical-cardiac and non-cardiac categories (**Table 1**). At the time of arrest, they were more likely to be receiving anti-arrhythmic infusions, continuous blood pressure monitoring, ECG monitoring, invasive assisted ventilation, and vasoactive agents, compared to medical-cardiac and non-cardiac patients. In surgical-cardiac patients, more cardiac arrests occurred in teaching hospitals, hospitals with more than 100 pediatric beds, and in hospitals with large pediatric and neonatal intensive care units.

**Table 1:** Pre-arrest and Hospital Characteristics by Illness Category.

| Table 1: Pre-arrest and Hospital Characteristics by Illness Category |  |  |  |  |
| --- | --- | --- | --- | --- |
|  | Illness Category |  |  |  |
|  | Surgical-Cardiac<br>(N = 3213) | Medical-Cardiac<br>(N = 3236) | Non-Cardiac<br>(N = 11247) | P-value |
| Age at event |  |  |  | <.001<sup>1</sup> |
| < 1 month | 1392 (43.3%) | 1165 (36.0%) | 3889 (34.6%) |  |
| 1 month - < 1 year | 1048 (32.6%) | 922 (28.5%) | 2928 (26.0%) |  |
| 1 year - < 8 years | 520 (16.2%) | 664 (20.5%) | 2503 (22.3%) |  |
| 8 years - < 18 years | 253 (7.9%) | 485 (15.0%) | 1927 (17.1%) |  |
| Sex |  |  |  | 0.465<sup>1</sup> |
| Male | 1747 (54.4%) | 1743 (53.9%) | 6196 (55.1%) |  |
| Female | 1466 (45.6%) | 1493 (46.1%) | 5052 (44.9%) |  |
| Location of event |  |  |  | <.001<sup>1</sup> |
| ICU | 2507 (78.0%) | 2438 (75.3%) | 8662 (77.0%) |  |
| ED | 16 (0.5%) | 247 (7.6%) | 710 (6.3%) |  |
| Floor/Ward | 141 (4.4%) | 186 (5.7%) | 898 (8.0%) |  |
| OR/PACU/Procedural Area | 345 (10.7%) | 260 (8.0%) | 793 (7.1%) |  |
| Other | 204 (6.3%) | 105 (3.2%) | 184 (1.6%) |  |
| Event witnessed | 3146 (97.9%) | 3063 (94.7%) | 10631 (94.5%) | <.001<sup>1</sup> |
| Night, weekend, or holiday event | 1344 (41.8%) | 1508 (46.6%) | 5502 (48.9%) | <.001<sup>1</sup> |
| **Pre-Existing Conditions** |  |  |  |  |
| Acute CNS non-stroke event | 51 (1.6%) | 135 (4.2%) | 805 (7.2%) | <.001<sup>1</sup> |
| Congestive heart failure (this admission or prior) | 910 (28.3%) | 909 (28.1%) | 333 (3.0%) | <.001<sup>1</sup> |
| Hypotension/Hypoperfusion | 1012 (31.5%) | 883 (27.3%) | 2921 (26.0%) | <.001<sup>1</sup> |
| Major trauma | 11 (0.3%) | 21 (0.7%) | 210 (1.9%) | <.001<sup>1</sup> |
| Metastatic or hematologic malignancy | 15 (0.5%) | 40 (1.2%) | 682 (6.1%) | <.001<sup>1</sup> |
| Metabolic/electrolyte abnormality | 448 (13.9%) | 509 (15.7%) | 2086 (18.5%) | <.001<sup>1</sup> |
| Myocardial ischemia/infarction (this admission or prior) | 35 (1.1%) | 62 (1.9%) | 44 (0.4%) | <.001<sup>1</sup> |
| Pneumonia | 58 (1.8%) | 165 (5.1%) | 1023 (9.1%) | <.001<sup>1</sup> |
| Renal insufficiency | 261 (8.1%) | 254 (7.8%) | 1168 (10.4%) | <.001<sup>1</sup> |
| Respiratory insufficiency | 1819 (56.6%) | 1915 (59.2%) | 8056 (71.6%) | <.001<sup>1</sup> |
| Sepsis/pneumonia (including septicemia) | 278 (8.6%) | 500 (15.4%) | 2794 (24.8%) | <.001<sup>1</sup> |
| **Interventions at Place at Time of Event** |  |  |  |  |
| Antiarrhythmic infusions in place at time of arrest | 112 (3.5%) | 82 (2.5%) | 46 (0.4%) | <.001<sup>1</sup> |
| Arterial catheter in place at time of arrest | 1581 (49.2%) | 713 (22.0%) | 2170 (19.3%) | <.001<sup>1</sup> |
| Dialysis in place at time of arrest | 90 (2.8%) | 30 (0.9%) | 291 (2.6%) | <.001<sup>1</sup> |
| ECG monitor in place at time of arrest | 3064 (95.4%) | 2848 (88.0%) | 9878 (87.8%) | <.001<sup>1</sup> |
| Supplemental oxygen in place at time of arrest | 758 (23.6%) | 820 (25.3%) | 2511 (22.3%) | 0.003<sup>1</sup> |
| Type of assisted ventilation |  |  |  | <.001<sup>1</sup> |
| None | 664 (20.7%) | 1013 (31.3%) | 2884 (25.6%) |  |
| Invasive assisted ventilation | 2270 (70.7%) | 1861 (57.5%) | 7087 (63.0%) |  |
| Non-invasive assisted ventilation | 279 (8.7%) | 362 (11.2%) | 1276 (11.3%) |  |
| Vascular access in place at time of arrest | 3025 (94.2%) | 2823 (87.2%) | 9826 (87.4%) | <.001<sup>1</sup> |
| Vasoactive agent in place at time of arrest | 1571 (48.9%) | 956 (29.5%) | 2803 (24.9%) | <.001<sup>1</sup> |
| **Hospital Characteristics** |  |  |  |  |
| Pediatric beds |  |  |  | <.001<sup>2</sup> |
| 0 - 50 | 386 (12.0%) | 839 (25.9%) | 2841 (25.3%) |  |
| 51 - 100 | 506 (15.8%) | 699 (21.6%) | 2745 (24.4%) |  |
| 101 - 200 | 1468 (45.7%) | 907 (28.0%) | 3440 (30.6%) |  |
| 201 + | 853 (26.5%) | 792 (24.5%) | 2222 (19.8%) |  |
| Pediatric intensive care beds |  |  |  | <.001<sup>2</sup> |
| 0 - 20 | 464 (14.4%) | 996 (30.8%) | 3752 (33.4%) |  |
| 21 - 40 | 1018 (31.7%) | 941 (29.1%) | 3227 (28.7%) |  |
| 41 + | 1731 (53.9%) | 1299 (40.1%) | 4268 (37.9%) |  |
| Neonatal intensive care beds |  |  |  | 0.371<sup>2</sup> |
| 0 - 30 | 1006 (31.3%) | 854 (26.4%) | 3130 (27.8%) |  |
| 31 - 50 | 734 (22.8%) | 1074 (33.2%) | 3514 (31.2%) |  |
| 51 + | 1473 (45.8%) | 1308 (40.4%) | 4604 (40.9%) |  |
| Teaching hospital | 3204 (99.7%) | 3180 (98.3%) | 11047 (98.2%) | <.001<sup>1</sup> |
| **Arrest Characteristics** |  |  |  |  |
| First documented rhythm <sup>3</sup> |  |  |  | <.001<sup>1</sup> |
| Bradycardia | 1783 (55.5%) | 1872 (57.8%) | 7063 (62.8%) |  |
| Asystole | 334 (10.4%) | 420 (13.0%) | 1660 (14.8%) |  |
| PEA | 671 (20.9%) | 577 (17.8%) | 1941 (17.3%) |  |
| Ventricular tachycardia | 83 (2.6%) | 118 (3.6%) | 196 (1.7%) |  |
| Ventricular fibrillation | 160 (5.0%) | 119 (3.7%) | 124 (1.1%) |  |
| Other | 182 (5.7%) | 131 (4.0%) | 264 (2.3%) |  |
| CPR duration (minutes, truncated after 120 minutes) | 12.0 (3.4, 34.0) | 13.0 (4.0, 32.3) | 9.3 (3.0, 25.0) | <.001<sup>2</sup> |
| ECPR | 655 (20.4%) | 288 (8.9%) | 336 (3.0%) | <.001<sup>1</sup> |
| Total number of epinephrine doses | 2.0 (1.0, 5.0) | 2.0 (1.0, 5.0) | 2.0 (0.0, 4.0) | <.001<sup>2</sup> |
| Time to first epinephrine (minutes, truncated after 30 minutes) <sup>4</sup> | 1.0 (0.0, 3.0) | 2.0 (0.0, 4.0) | 1.9 (0.0, 3.6) | <.001<sup>2</sup> |
| **Pharmacological Interventions During Event** |  |  |  |  |
| Vasopressor(s) other than epinephrine bolus | 1308 (40.7%) | 913 (28.2%) | 2817 (25.0%) | <.001<sup>1</sup> |
| Epinephrine bolus administered | 2649 (82.5%) | 2458 (76.0%) | 7864 (69.9%) | <.001<sup>1</sup> |
| Sodium bicarbonate administered | 1600 (49.8%) | 1452 (44.9%) | 3855 (34.3%) | <.001<sup>1</sup> |
| Atropine administered | 517 (16.1%) | 558 (17.2%) | 2013 (17.9%) | 0.056<sup>1</sup> |
| Amiodarone administered | 176 (5.5%) | 171 (5.3%) | 181 (1.6%) | <.001<sup>1</sup> |
| Calcium chloride or gluconate administered | 1533 (47.7%) | 1156 (35.7%) | 2881 (25.6%) | <.001<sup>1</sup> |
| Lidocaine administered | 202 (6.3%) | 197 (6.1%) | 259 (2.3%) | <.001<sup>1</sup> |

<sup>1</sup> Likelihood ratio test. <sup>2</sup> Kruskal-Wallis test.
<sup>3</sup> First documented rhythm other includes neonatal VT and VF.
<sup>4</sup> Time to first epinephrine calculated for those with at least one dose of epinephrine.

Medical-cardiac patients were more likely to have an arrest in the emergency department and were less likely to have dialysis in place at time of arrest compared to surgical-cardiac and non-cardiac illness category patients.

Non-cardiac illness category patients were more likely to have pre-existing conditions of acute non-stroke central nervous system dysfunction, malignancy, trauma, respiratory insufficiency, and sepsis/pneumonia and were less likely to have congestive heart failure or myocardial ischemia than both surgical-cardiac and medical-cardiac illness categories.

### Arrest characteristics based on illness category

Patients with surgical- and medical-cardiac disease had more ventricular tachycardia or ventricular fibrillation as the first documented rhythm, longer durations of CPR and received more doses of epinephrine than non-cardiac patients (**Table 1**). Subjects with non-cardiac disease had higher rates of bradycardia as the first identified rhythm. The median time to first epinephrine dose administration was shortest in the surgical-cardiac patients and surgical-cardiac patients were more likely to receive epinephrine boluses, bolus doses of vasopressors other than epinephrine, sodium bicarbonate, and calcium than the medical-cardiac and non-cardiac groups. Non-cardiac patients received less lidocaine and amiodarone than surgical-cardiac and medical- cardiac patients.

### Outcomes based on illness category

Surgical-cardiac patients had the highest rate of event survival and survival to hospital discharge compared to medical-cardiac and non-cardiac, p<0.001 (**Table 2**). Surgical-cardiac and medical-cardiac patients had lower rates of ROSC (65% and 66%, respectively), compared to non-cardiac patients (72%). Rates of ROC with ECMO support were highest in surgical-cardiac patients (19.9%) and lower in medical-cardiac (8.5%) and non-cardiac patients (2.5%), p<0.001.

**Table 2:**
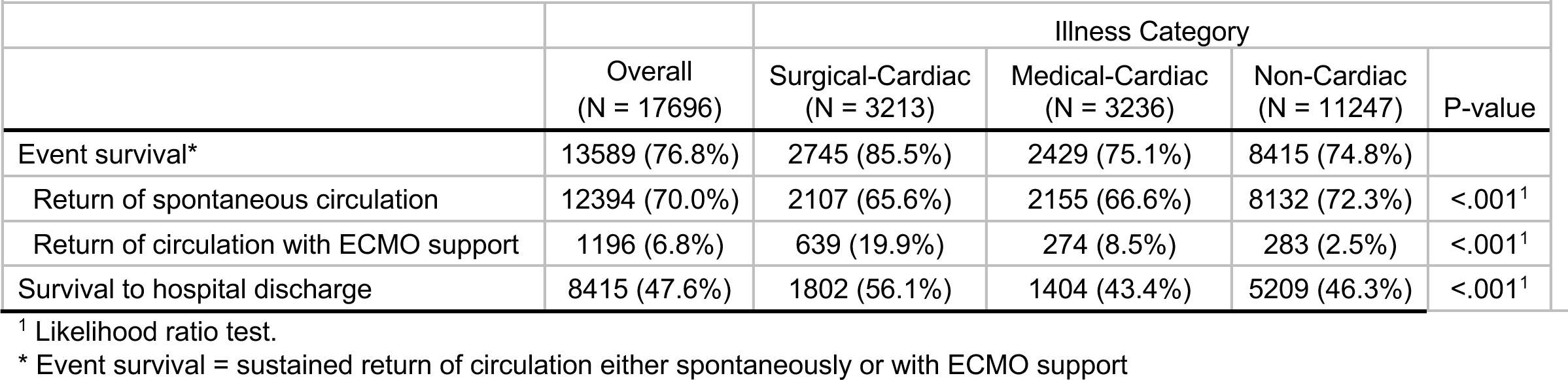
Outcomes by Illness Category.

| **Table 2: Outcomes by Illness Category** |  |  |  |  |  |
| --- | --- | --- | --- | --- | --- |
|  |  | Illness Category |  |  |  |
|  | Overall<br>(N = 17696) | Surgical-Cardiac<br>(N = 3213) | Medical-Cardiac<br>(N = 3236) | Non-Cardiac<br>(N = 11247) | P-value |
| Event survival* | 13589 (76.8%) | 2745 (85.5%) | 2429 (75.1%) | 8415 (74.8%) |  |
| Return of spontaneous circulation | 12394 (70.0%) | 2107 (65.6%) | 2155 (66.6%) | 8132 (72.3%) | <.001<sup>1</sup> |
| Return of circulation with ECMO support | 1196 (6.8%) | 639 (19.9%) | 274 (8.5%) | 283 (2.5%) | <.001<sup>1</sup> |
| Survival to hospital discharge | 8415 (47.6%) | 1802 (56.1%) | 1404 (43.4%) | 5209 (46.3%) | <.001<sup>1</sup> |

<sup>1</sup> Likelihood ratio test.
\* Event survival = sustained return of circulation either spontaneously or with ECMO support

When adjusted for age at arrest, location of event, and hospital size, and using non-cardiac illness category patients as reference, surgical-cardiac patients had the highest odds of survival to hospital discharge (aOR 1.28, [95% CI 1.16, 1.40], p<0.001) and medical-cardiac patients had the lowest odds of survival (aOR 0.87, [95% CI 0.80, 0.95], p<0.001) (**Table 3**).

**Table 3:** Multivariable Mixed Logistic Regression on Survival to Hospital Discharge.

|  |  |  |
| --- | --- | --- |
|  | Survival to hospital discharge<br>Odds ratio (95% CI) | P-value |
| **Age at CPA event** |  | <.001 |
| < 1 month | Reference |  |
| 1 month - < 1 year | 1.97 (1.82, 2.14) |  |
| 1 year - < 8 years | 1.16 (1.07, 1.27) |  |
| 8 years - < 18 years | 0.66 (0.60, 0.73) |  |
| **Location of event** |  | <.001 |
| ICU | Reference |  |
| ED | 0.74 (0.64, 0.86) |  |
| Floor/Ward | 1.91 (1.69, 2.17) |  |
| OR/PACU/Procedural Area | 2.82 (2.49, 3.20) |  |
| Other | 1.30 (1.07, 1.58) |  |
| **Hospital size (number of pediatric beds)** |  | 0.264 |
| 0 - 50 | Reference |  |
| 51 - 100 | 1.02 (0.84, 1.25) |  |
| 101 - 200 | 1.19 (0.93, 1.52) |  |
| 201 + | 1.29 (0.93, 1.81) |  |
| **Illness category** |  | <.001 |
| Surgical-Cardiac | 1.28 (1.16, 1.40) |  |
| Medical-Cardiac | 0.87 (0.80, 0.95) |  |
| Non-Cardiac | Reference |  |

Results are based on a multivariable model(s), adjusting for each of the predictors in this table. Model(s) account for correlated observations.

### Outcome and management trends over time by illness categories

Rates of survival to hospital discharge significantly increased over time for all three categories: surgical-cardiac from 45.6% in 2000-2004 to 62.3% in 2015-2021 (OR 1.24), medical-cardiac from 36.6% to 47.0% (OR 1.14), and non-cardiac from 34.3% to 52.0% (OR 1.22), (all p<0.001) (**Table 4**, **Figure 2**). Event survival also increased for all groups over time. While rates of ROSC did not statistically improve over time for the surgical-cardiac group (56.7% in 2000-2004 to 65.2% in 2015-2021; OR=1.04, p=0.327), event survival rate increased due to increase in ROC with ECMO (a 31% increase of odds of ROC with ECMO for each 5- year era). ROSC increased over time for medical-cardiac (OR 1.09; p=0.021) and non-cardiac patients (OR 1.10; p<0.001). ROC with ECMO increased for both medical-cardiac and non-cardiac patients with 42% and 47% increase of odds for each 5-year era, respectively (p<0.001).

**Table 4:**
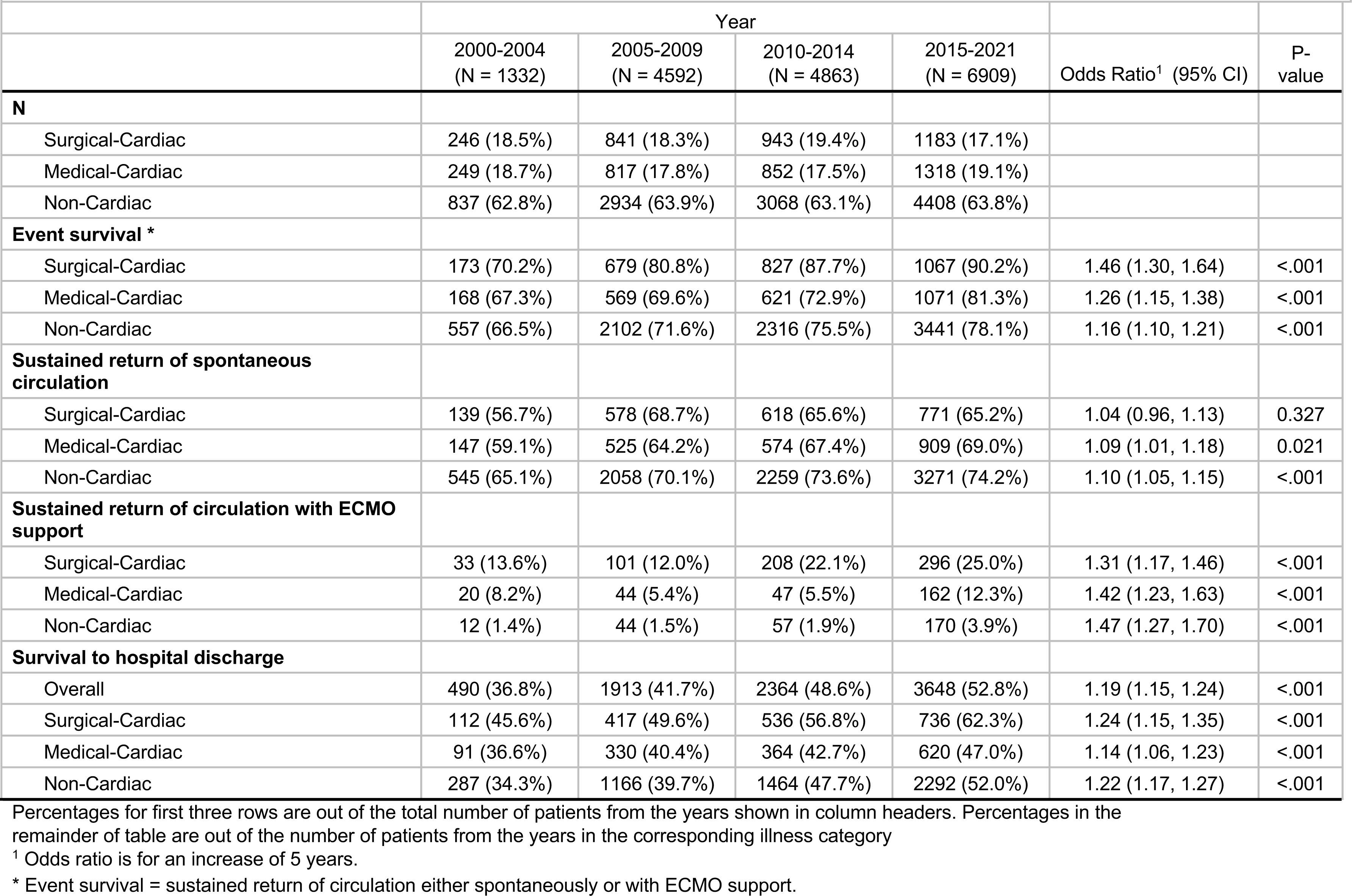
Trends in Cardiac Arrest Outcomes by Time.

|  | Year |  |  |  |  |  |
| --- | --- | --- | --- | --- | --- | --- |
|  | 2000-2004<br>(N = 1332) | 2005-2009<br>(N = 4592) | 2010-2014<br>(N = 4863) | 2015-2021<br>(N = 6909) | Odds Ratio<sup>1</sup> (95% CI) | P-value |
| **N** |  |  |  |  |  |  |
| Surgical-Cardiac | 246 (18.5%) | 841 (18.3%) | 943 (19.4%) | 1183 (17.1%) |  |  |
| Medical-Cardiac | 249 (18.7%) | 817 (17.8%) | 852 (17.5%) | 1318 (19.1%) |  |  |
| Non-Cardiac | 837 (62.8%) | 2934 (63.9%) | 3068 (63.1%) | 4408 (63.8%) |  |  |
| **Event survival \*** |  |  |  |  |  |  |
| Surgical-Cardiac | 173 (70.2%) | 679 (80.8%) | 827 (87.7%) | 1067 (90.2%) | 1.46 (1.30, 1.64) | <.001 |
| Medical-Cardiac | 168 (67.3%) | 569 (69.6%) | 621 (72.9%) | 1071 (81.3%) | 1.26 (1.15, 1.38) | <.001 |
| Non-Cardiac | 557 (66.5%) | 2102 (71.6%) | 2316 (75.5%) | 3441 (78.1%) | 1.16 (1.10, 1.21) | <.001 |
| **Sustained return of spontaneous circulation** |  |  |  |  |  |  |
| Surgical-Cardiac | 139 (56.7%) | 578 (68.7%) | 618 (65.6%) | 771 (65.2%) | 1.04 (0.96, 1.13) | 0.327 |
| Medical-Cardiac | 147 (59.1%) | 525 (64.2%) | 574 (67.4%) | 909 (69.0%) | 1.09 (1.01, 1.18) | 0.021 |
| Non-Cardiac | 545 (65.1%) | 2058 (70.1%) | 2259 (73.6%) | 3271 (74.2%) | 1.10 (1.05, 1.15) | <.001 |
| **Sustained return of circulation with ECMO support** |  |  |  |  |  |  |
| Surgical-Cardiac | 33 (13.6%) | 101 (12.0%) | 208 (22.1%) | 296 (25.0%) | 1.31 (1.17, 1.46) | <.001 |
| Medical-Cardiac | 20 (8.2%) | 44 (5.4%) | 47 (5.5%) | 162 (12.3%) | 1.42 (1.23, 1.63) | <.001 |
| Non-Cardiac | 12 (1.4%) | 44 (1.5%) | 57 (1.9%) | 170 (3.9%) | 1.47 (1.27, 1.70) | <.001 |
| **Survival to hospital discharge** |  |  |  |  |  |  |
| Overall | 490 (36.8%) | 1913 (41.7%) | 2364 (48.6%) | 3648 (52.8%) | 1.19 (1.15, 1.24) | <.001 |
| Surgical-Cardiac | 112 (45.6%) | 417 (49.6%) | 536 (56.8%) | 736 (62.3%) | 1.24 (1.15, 1.35) | <.001 |
| Medical-Cardiac | 91 (36.6%) | 330 (40.4%) | 364 (42.7%) | 620 (47.0%) | 1.14 (1.06, 1.23) | <.001 |
| Non-Cardiac | 287 (34.3%) | 1166 (39.7%) | 1464 (47.7%) | 2292 (52.0%) | 1.22 (1.17, 1.27) | <.001 |

Percentages for first three rows are out of the total number of patients from the years shown in column headers. Percentages in the remainder of table are out of the number of patients from the years in the corresponding illness category
<sup>1</sup> Odds ratio is for an increase of 5 years.
\* Event survival = sustained return of circulation either spontaneously or with ECMO support.

**Figure 2.**
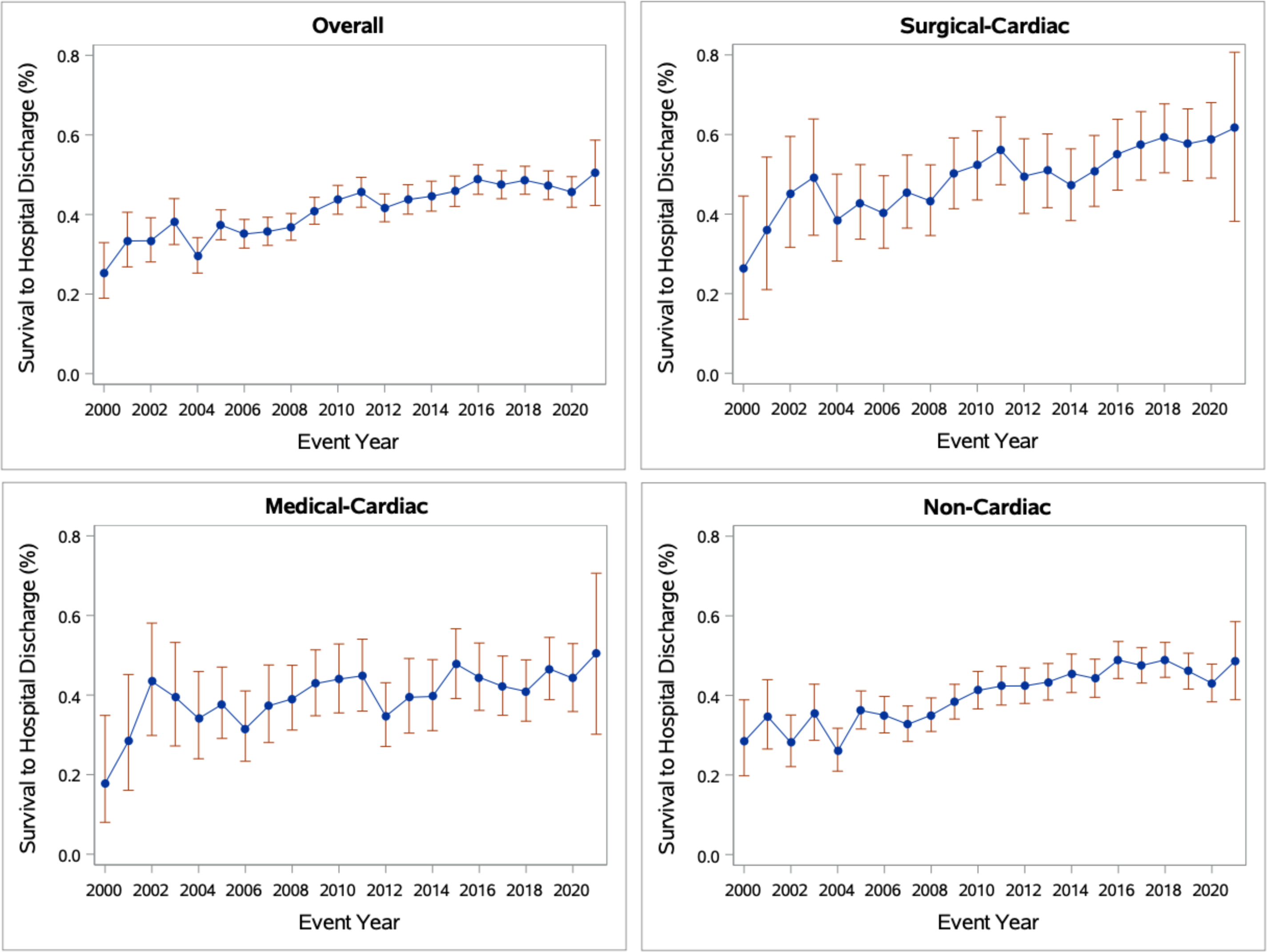
Trend of survival to hospital discharge of overall cohort and based on illness category over time.

Rates of ECPR significantly increased over time periods for all categories (**Table 5**). ECPR rates were highest in the surgical-cardiac patients and increased from 16.5% to 25.4% (28% increased adjusted odds of ECPR per 5-year era). ECPR rate in medical-cardiac patients increased from 9% in the 2000-2004 era, to 12.7% in the 2015-2021: an increased odds of ECPR 39% for every era. Non-cardiac patients had the lowest rates of ECPR although we also observed an increase in ECPR utilization from 1.8% to 4.6% between the first and last eras, corresponding to 44% increased odds with every era, the greatest rate of increase among all the groups (all p<0.001).

**Table 5:** Trends in Cardiac Arrest Management by Time.

| Table 5: Trends in Cardiac Arrest Management by Time |  |  |  |  |  |  |  |
| --- | --- | --- | --- | --- | --- | --- | --- |
|  | Year |  |  |  |  |  |  |
|  | 2000-2004<br>(N = 1332) | 2005-2009<br>(N = 4592) | 2010-2014<br>(N = 4863) | 2015-2021<br>(N = 6909) | Odds Ratio<sup>1</sup><br>(95% CI) | Effect<sup>1</sup> (95% CI) | P-value |
| **N** |  |  |  |  |  |  |  |
| Surgical-Cardiac | 246 (18.5%) | 841 (18.3%) | 943 (19.4%) | 1183 (17.1%) |  |  |  |
| Medical-Cardiac | 249 (18.7%) | 817 (17.8%) | 852 (17.5%) | 1318 (19.1%) |  |  |  |
| Non-Cardiac | 837 (62.8%) | 2934 (63.9%) | 3068 (63.1%) | 4408 (63.8%) |  |  |  |
| **ECPR** |  |  |  |  |  |  |  |
| Surgical-Cardiac | 40 (16.5%) | 102 (12.1%) | 212 (22.4%) | 301 (25.4%) | 1.28 (1.15, 1.42) |  | <.001 |
| Medical-Cardiac | 22 (9.0%) | 47 (5.8%) | 51 (6.0%) | 168 (12.7%) | 1.39 (1.22, 1.60) |  | <.001 |
| Non-Cardiac | 15 (1.8%) | 52 (1.8%) | 67 (2.2%) | 202 (4.6%) | 1.44 (1.26, 1.66) |  | <.001<sup>2</sup> |
| **CPR duration (minutes, truncated after 120 minutes)** |  |  |  |  |  |  |  |
| Surgical-Cardiac | 19.6 (6.6, 44.9) | 14.8 (5.0, 35.1) | 10.9 (3.0, 31.8) | 9.0 (3.0, 32.2) |  | -2.24 (-3.20, -1.27) | <.001 |
| Medical-Cardiac | 24.5 (11.0, 44.6) | 15.1 (5.0, 33.3) | 12.1 (4.0, 30.9) | 9.7 (3.0, 30.0) |  | -2.49 (-3.39, -1.59) | <.001 |
| Non-Cardiac | 18.9 (7.6, 37.0) | 11.2 (4.0, 26.0) | 8.8 (3.0, 23.8) | 7.0 (2.0, 23.0) |  | -2.31 (-2.76, -1.85) | <.001 |
| **Total number of epinephrine doses** |  |  |  |  |  |  |  |
| Surgical-Cardiac | 2.1 (1.0, 5.2) | 2.0 (1.0, 5.0) | 2.0 (1.0, 5.0) | 2.0 (1.0, 6.0) |  | 0.41 (0.22, 0.59) | <.001 |
| Medical-Cardiac | 3.0 (1.0, 5.0) | 2.0 (1.0, 4.7) | 2.0 (0.0, 4.6) | 2.0 (0.3, 5.0) |  | 0.17 (0.02, 0.33) | 0.031 |
| Non-Cardiac | 2.1 (1.0, 5.0) | 2.0 (0.0, 4.0) | 1.0 (0.0, 4.0) | 1.0 (0.0, 4.0) |  | -0.01 (-0.09, 0.07) | 0.774 |
| **Time to first epinephrine (minutes, truncated after 30 minutes)<sup>3</sup>** |  |  |  |  |  |  |  |
| Surgical-Cardiac | 1.3 (0.0, 3.7) | 1.0 (0.0, 3.5) | 1.0 (0.0, 3.0) | 1.0 (0.0, 2.0) |  | -0.36 (-0.48, -0.23) | <.001 |
| Medical-Cardiac | 1.2 (0.0, 5.0) | 2.0 (0.0, 4.2) | 2.0 (0.0, 4.0) | 2.0 (0.0, 3.0) |  | -0.29 (-0.45, -0.13) | <.001 |
| Non-Cardiac | 1.8 (0.0, 4.8) | 2.0 (0.0, 4.0) | 1.4 (0.0, 3.4) | 1.3 (0.0, 3.0) |  | -0.22 (-0.31, -0.13) | <.001 |
| **Sodium bicarbonate administered during event** |  |  |  |  |  |  |  |
| Surgical-Cardiac | 159 (64.9%) | 470 (55.8%) | 432 (45.8%) | 539 (45.6%) | 0.76 (0.70, 0.83) |  | <.001 |
| Medical-Cardiac | 172 (68.8%) | 413 (50.6%) | 343 (40.3%) | 524 (39.8%) | 0.76 (0.70, 0.82) |  | <.001 |
| Non-Cardiac | 455 (54.4%) | 1184 (40.4%) | 951 (31.0%) | 1265 (28.7%) | 0.76 (0.72, 0.79) |  | <.001 |
| **Atropine administered during event** |  |  |  |  |  |  |  |
| Surgical-Cardiac | 74 (30.2%) | 203 (24.1%) | 150 (15.9%) | 90 (7.6%) | 0.56 (0.50, 0.63) |  | <.001 |
| Medical-Cardiac | 100 (40.2%) | 202 (24.7%) | 142 (16.7%) | 114 (8.6%) | 0.52 (0.47, 0.58) |  | <.001 |
| Non-Cardiac | 336 (40.1%) | 669 (22.8%) | 571 (18.6%) | 437 (9.9%) | 0.54 (0.51, 0.58) |  | <.001 |
| **Calcium chloride or gluconate administered during event** |  |  |  |  |  |  |  |
| Surgical-Cardiac | 140 (57.0%) | 424 (50.5%) | 409 (43.4%) | 559 (47.2%) | 0.89 (0.83, 0.97) |  | 0.006 |
| Medical-Cardiac | 129 (51.9%) | 298 (36.4%) | 274 (32.2%) | 454 (34.5%) | 0.88 (0.81, 0.94) |  | <.001 |
| Non-Cardiac | 346 (41.4%) | 807 (27.5%) | 702 (22.9%) | 1025 (23.3%) | 0.83 (0.79, 0.87) |  | <.001 |

Percentages for first three rows are out of the total number of patients from the years shown in column headers. Percentages in the remainder of table are out of the number of patients from the years in the corresponding illness category
<sup>1</sup> Odds ratio and effect is for an increase of 5 years.
<sup>2</sup> p-value for ECPR calculated for sites with at least 5 subjects who had ECPR.
<sup>3</sup> Time to first epinephrine calculated for those with at least one dose of epinephrine.

CPR duration and time to first epinephrine dose were shorter over time for all three illness categories, and rates of administration of sodium bicarbonate, atropine and calcium decreased over time.

## DISCUSSION

Our retrospective analysis from the GWTG®-R registry shows the rate of survival to hospital discharge following IHCA for all pediatric patients with cardiac and non-cardiac disease has increased from 37% to 53% over the prior two decades. Overall, patients with surgical-cardiac disease had the highest rates of survival to hospital discharge, while medical-cardiac patients had the lowest rates of survival, compared to the non-cardiac group. Although survival improved over the 20-year observation period across all three illness categories, the increase was largest in surgical-cardiac patients. Additionally, the use of ECPR increased in all three illness categories over time, contributing to the higher rates of event survival due to higher of return of circulation on ECMO.

Survival to hospital discharge from pediatric IHCA improved across cardiac and non-cardiac illness categories over time, with highest rates in the 2015-2021 epoch. Holmberg *et al.* reported survival to hospital discharge in 2018 of 38% for pulseless events and 66% for non- pulseless events, describing a plateau in survival in 2010^4^. We did not identify a plateau in 2015 but instead saw increasing rates of survival across all epochs. This may be because we specifically focused on three distinct illness categories and excluded others such as trauma and delivery room resuscitations that were included in the analyses by Holmberg and colleagues^14^. Additionally, we did not perform our analyses based on first documented rhythm, but instead by illness category; and patients with non-cardiac illness categories had more bradycardia than surgical- and medical-cardiac patients.

The non-cardiac patients had higher odds of survival to hospital discharge than medical- cardiac patients, though the highest rates of survival occurred in the surgical-cardiac patients. These populations are different for several reasons. Surgical-cardiac patients’ cause of arrest is usually related to hypotension, possibly due to transient post-operative low cardiac output or from surgical residual disease, and thus arrest care can focus on restoring sufficient coronary perfusion and blood pressure to obtain return of spontaneous circulation^5^. When ROSC is not possible, our data show higher rates of ECPR utilization in this patient population, consistent with other studies and AHA recommendations for this population if resources are available^26^. The medical-cardiac category, on the other hand, may be multi-factorial in nature, where resuscitation must include management of cardiac, respiratory, and other organ involvement. This may contribute to lower rates of arrest survival, a disparity which was also found in a recent publication from the Pediatric Cardiac Critical Care Consortium (PC^4^)^3^. Non-cardiac patients had higher rates of respiratory insufficiency and more frequently had bradycardia with poor perfusion as a first documented rhythm^27^, all of which have been associated with higher rates of survival . Our findings differ from Ortmann *et al*. who found that survival to hospital to discharge rates were higher for medical-cardiac than non-cardiac patients, though our findings are similar in that surgical-cardiac outcomes were better than all other categories^14, 15^. This difference may be explained by differences in our initial cohort. We excluded trauma illness category patients from our non-cardiac group, a group that traditionally has particularly poor outcomes. Furthermore, Ortmann *et al.* excluded the newborn illness category, whereas we imputed the illness category in these patients prior to the newborn illness category’s removal in 2016 to avoid temporal biases in exclusion criteria.

Consistent with previously reported data in both adults^28–32^ and children^2^, we found an increased rate of survival to hospital discharge over the past 20 years. Specific to our analysis, the increased rate of survival to hospital discharge was found across all illness categories. This may be for several reasons, and attributable to the advancement of resuscitation science, with a focus on early recognition, high-quality CPR, and diligent post-cardiac arrest care^33^. Importantly, we found ECPR utilization increased across all categories (as has been seen in other adult and pediatric studies^34, 35^), particularly in the post-operative surgical-cardiac population^36^. This contributes to higher rates of event survival with return of circulation with ECMO support, and there was also a smaller contribution from increasing ROSC across the eras in all illness categories (and non-significant in the medical-cardiac patients). ECMO is more commonly used in large centers with systems in place to support the advanced technology: surgeons, ECMO specialists, protocols, equipment, and expertise. In 2020 the AHA stated that “ECMO may be considered for pediatric patients with cardiac diagnoses who have an IHCA….”^37^. Due to lack of data, there was no guideline specific to the non-cardiac population. Despite this lack of guideline-specific utilization of ECPR in non-cardiac patients, the greatest increase occurred in the non-cardiac illness category with a 44% increase for every 5-year era. This may reflect changes in the practice of ECMO use in the growingly complex subject category without cardiac disease.

Over the last two decades the timing and rates of administration of medications during CPR such as epinephrine, calcium, bicarbonate, and atropine have changed across all three categories, aligning more in agreement with AHA PALS Guidelines^37^. Across all categories, time to first dose of epinephrine decreased on average with every 5-year era. Delay to epinephrine dosing in prior GWTG®-R work was associated with lower rates of ROSC, and survival to hospital discharge^38^, and thus the reduction in time to first dose of epinephrine over time, though small, may be an important factor associated with higher rates of ROSC and survival seen across the categories^26^. Beyond epinephrine, the analysis of pharmacologic interventions during cardiac arrest showed the use of sodium bicarbonate, atropine, and calcium has decreased over the study period in all categories, though remained highest in the surgical-cardiac category. Sodium bicarbonate and calcium carbonate have been consistently associated with lower rates of survival in the general pediatric and cardiac populations, and perhaps continued reduction in this practice will lead to even better outcomes in the future^26, 39–43^.

Our findings beg the question as to whether the disparities among the illness categories reflect the patients, or the systems in which they are cared for. How can we narrow the gap in the outcomes of these patient categories? In large centers, ECMO utilization may be “spilling” over from the post-surgical cardiac patients to medical-cardiac and non-cardiac patients which may improve survival through obtaining return of circulation and treating the underlying cause of arrest. Furthermore, larger investigations of the cardiac population, specifically the medical- cardiac population, and better understanding of physiology-directed CPR (end-tidal CO^2^, diastolic blood pressure, cerebral near infrared spectroscopy) and post-cardiac arrest care may continue to improve outcomes and lessen the disparities.

As with any study using a large multicenter registry, our study does have limitations. GWTG®-R has a robust process to ensure data integrity and validation as there are several sites that submit data for the registry, thus limiting variability. There is still missing data across the cohort, at times excluding variables with high levels of missingness (>20%) that could limit analyses. We utilized imputation for these variables, particularly in the newborn illness category which could bias the results; however, our findings of similar outcomes across illness categories from other reports is suggestive that this imputation provided similar groups for comparison. GWTG®-R is a voluntary registry, and thus may not be inclusive of all centers providing pediatric care across the United States. Additionally, centers may submit data during different time periods over our period of observation. We utilized a mixed-effects model to consider center-level variability of data submission but cannot fully control for differences in post-arrest care.

## CONCLUSION

Over the last 20 years, both survival to hospital discharge and ECPR use have increased in all IHCA illness categories. Children with surgical-cardiac IHCA have higher odds of survival to hospital discharge compared to non-cardiac IHCA categories, whereas odds of survival were worst with medical-cardiac IHCA.

## Data Availability

Raw data were generated via AHA GWTG. Derived data supporting the findings of this study are available from the corresponding author MMG on request.

## AKNOWLEDGEMENTS

The Get With The Guidelines^®^-Resuscitation program is provided by the American Heart Association. We recognize the American Heart Association’s Get With the Guidelines^®^- Resuscitation Pediatric Research Task Force members: Anne-Marie Guerguerian MD PhD FRCPC FAAP FAHA; Caitlin E. O’Brien MD MPH; Ericka L. Fink MD MS; Javier J. Lasa MD FAAP; Joan S. Roberts MD; Lillian Su MD; Linda L. Brown MD MSCE; Maya Dewan MD MPH; Melania M. Bembea MD MPH PhD; Monica Kleinman MD; Noorjahan Ali MD MS FAAP; Punkaj Gupta MBBS; Robert M. Sutton MD MSCE; Ron Reeder MS PhD; and Todd Sweberg MD MBA.

